# Gait parameters, Imaging features, and CSF tap test in idiopathic Normal Pressure Hydrocephalus (iNPH) Is composite testing the way to go?

**DOI:** 10.1101/2025.11.16.25340322

**Authors:** Sagar Poudel, Alfonso Fasano, Deepa Dash, Aparna Wagle Shukla, Ajay Garg, Ashish Dutt Upadhyay, Naveet Wig, Roopa Rajan, Animesh Das, MR Divya, Manjari Tripathi, Achal K. Srivastava, SS Kale, P Sarat Chandra, Ashish Suri, Pramod K Pal, Hrishikesh Kumar, Deepti Vibha, Rajesh Kumar Singh, Jasmine Parihar, Sakoon Saggu, Ranveer Singh Jadon, Ved Prakash Meena, Bindu Prakash, Arunmozhimaran Elavarasi

## Abstract

**Introduction:** Idiopathic normal pressure hydrocephalus is characterized by gait disturbance, cognitive decline, and urinary dysfunction which may improve with ventriculo-peritoneal shunting. This study evaluated the role of conventional MRI features and the CSF tap test (CSF-TT) in predicting shunt responsiveness in iNPH.

**Methods:** This is an Ambispective cohort study of 40 patients with probable iNPH evaluated between 2019 and 2024. Baseline MRI parameters, gait features, iNPH score, CSF opening pressure were analyzed. Functional outcome was assessed using the modified Rankin scale (mRS) at baseline, 24 hours after CSF-TT, and 24 weeks after VP shunt surgery. CSF-TT responders were defined as at least a 1-point improvement in mRS 24 hours after CSF-TT. The diagnostic performance of individual MRI parameters and composite diagnostic parameters were evaluated.

**Results:** Forty patients underwent CSF-TT. There were no significant differences between CSF-TT responders and non-responders in the baseline clinical gait parameters, the iNPH scale, and MRI findings. Turning disturbance, wide-based stride, and reduced foot clearance showed significant improvement after CSF-TT. Individual MRI parameters and CSF-TT parameters showed limited value in predicting shunt responsiveness. Composite diagnostic criteria combining CSF-TT and CSF opening pressure >18cm H20 showed sensitivity of 62.5 % and specificity of 71.4% with the highest Youden index indicating modest diagnostic accuracy in predicting shunt responsiveness.

**Conclusion:** Shunt surgery provided significant functional benefit in iNPH. Neither the CSF tap test nor conventional MRI markers alone reliably predicted shunt responsiveness. A multimodal assessment combining imaging, clinical evaluation, and CSF dynamics is required to optimize patient selection.

## Introduction

Idiopathic Normal Pressure Hydrocephalus is a condition characterized by a triad of gait disturbance, urinary disturbance, and cognitive impairment, and is treated by ventriculoperitoneal shunting.^1^ iNPH is suspected in the right clinical setting, when imaging suggests ventriculomegaly characterized by an Evans ratio >0.3^2^ or disproportionately enlarged subarachnoid space hydrocephalus (DESH), which are considered highly sensitive and specific for NPH.^3^ In the SINPHONI2 randomized trial, it was found that only 65% of patients with clinico-radiologic iNPH showed improvement following shunt surgery.^4^ Patients with imaging evidence of NPH often undergo CSF tap test before surgery, and several centers use the CSF-TT to select patients for VP shunt insertion.^5^ However, recent meta-analysis revealed that the diagnostic accuracy of CSF-TT was found to be suboptimal and not predictive of postoperative outcomes.^6^ It is not known which baseline parameter, score, or MRI finding correlates best with CSF-TT responsiveness. This suggests that currently available tools are not accurate enough to select patients who would benefit from surgery. In this paper, we analyzed the positive predictive values of specific imaging findings for CSF-TT responsiveness and shunt responsiveness. We also studied the parameters of gait that were associated with CSF tap test responsiveness. The diagnostic accuracy of CSF-TT in predicting shunt responsiveness^7^ and the short-term outcomes of shunt surgery^8^ in this cohort have been reported elsewhere. In our previous publication from this cohort, we found that CSF opening pressures could be a predictor of tap test responsiveness.^9^ We also did an exploratory analysis of whether composite diagnostic models incorporating CSF-TT and CSF opening pressures would perform better in predicting shunt responsiveness.

## Methods

In this Ambispective cohort study, Patients with at least one clinical feature of the triad, as per Relkin guidelines, underwent Magnetic resonance imaging of the brain and were subjected to a CSF tap test as per the Institutional protocol.^7^ Patients underwent assessment by evaluation of Boon’s gait score, iNPH score, Modified Rankin score, TUG test, and MoCA scores before and 24 hours after 30-50 ml of CSF-TT. Patients with at least 1 point improvement in the modified Rankin score 24 hours after CSF tap test were considered as tap test responders. However, patients were referred for the insertion of a programmable Ventriculoperitoneal shunt, regardless of CSF TT results. Shunt responders are defined as patients who show at least 1 point improvement in mRS after VP shunt surgery. The study was designed and reported in accordance with the STROBE reporting guidelines and was approved by the Institutional Review Board.

The case definitions of MRI findings applied in this study are summarized in Box 1 and representative images depicted in Figure 1.

**Figure 1.**
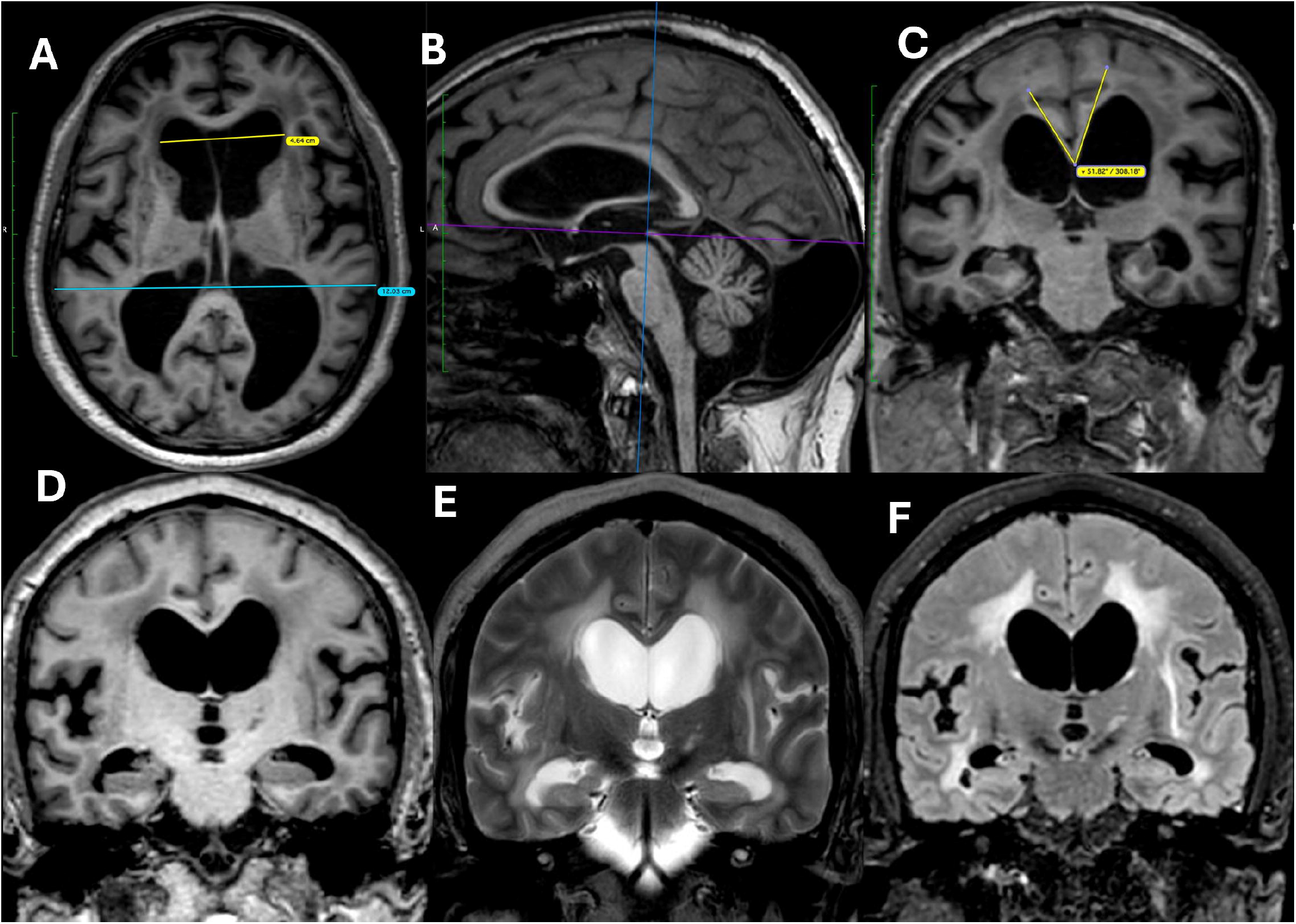
Axial T1-weighted image (A) shows ventriculomegaly out of proportion to cortical sulcal dilatation, with an Evans’ index of 0.39. Sagittal T1-weighted image (B) demonstrates the reference planes for callosal angle measurement: the horizontal line passing through the anterior and posterior commissures (AC–PC line) and the vertical line perpendicular to it, passing through the posterior commissure. Coronal T1-weighted image (C), corresponding to the vertical reference plane, shows a narrowed callosal angle of 52°, flattened high-convexity sulci, periventricular hypointensities, and widened Sylvian fissures, suggestive of **idiopathic normal pressure hydrocephalus (iNPH)**. Coronal T1 (D), T2 (E), and FLAIR (F) images demonstrate periventricular T2/FLAIR hyperintensities, a narrow callosal angle, and flattened convexity sulci.

### Box 1: Definitions of MRI findings

- **Periventricular white matter changes**^**10**^

Defined as areas of T2/FLAIR hyperintensity immediately adjacent to the lateral ventricles.

- **Bowing of the Corpus Callosum**^**2**^

Anterior and superior displacement of the corpus callosum body in a mid-sagittal section.

- **Evans’ Ratio**^**3**^

The maximum width of the frontal horn to the maximum inner skull diameter at the same axial slice.

- **Callosal angle**^**10**^

Measured on a coronal plane through the posterior commissure, perpendicular to the AC-PC line, as the angle formed between the medial wall of the lateral ventricles.

- **Infarcts**^**2**^

Defined as focal parenchymal lesions conforming to a vascular territory.

### Increased CSF opening pressure

Based on our previous analysis, we considered a cutoff of 18 cm of H2O to define higher CSF pressure within the NPH cohort.

## Statistical analysis

Data were collected and managed using REDCap (Research Electronic Data Capture), a secure, web-based application designed to support data capture for research studies hosted by the All India Institute of Medical Sciences (AIIMS), New Delhi, India. Data were exported and cleaned using Microsoft Excel (Microsoft Corp., Redmond, WA, USA). Descriptive statistics, including frequencies and percentages, were used to describe categorical variables. Mean and standard deviations were used for continuous variables with a normal distribution, while medians and interquartile ranges were used for non-parametric data. For comparisons between groups, an independent samples t-test was used for normally distributed data, and the Mann–Whitney U test was used for non-normally distributed data. Dichotomous variables were compared using the Chi-squared test or Fisher’s exact test. All statistical analyses were performed using Stata version 17 (StataCorp, College Station, TX, USA). A p-value of <0.05 was considered statistically significant.

## Results

### Patient Characteristics

Of 48 screened patients, 40 met criteria for probable iNPH and underwent a CSF-TT; 18 (45%) were responders. Twenty-four patients proceeded to shunt surgery, of whom 15 (62.5%) improved postoperatively. The duration of symptoms was significantly shorter among responders compared to non-responders (median, 25 vs. 31 months; p = 0.05). Baseline demographic and clinical characteristics were similar between CSF-TT responders and non-responders (Table 1). The age and sex distributions, as well as the presence of Parkinsonism, falls, and urinary/cognitive symptoms, were comparable between groups.

**Table 1:**
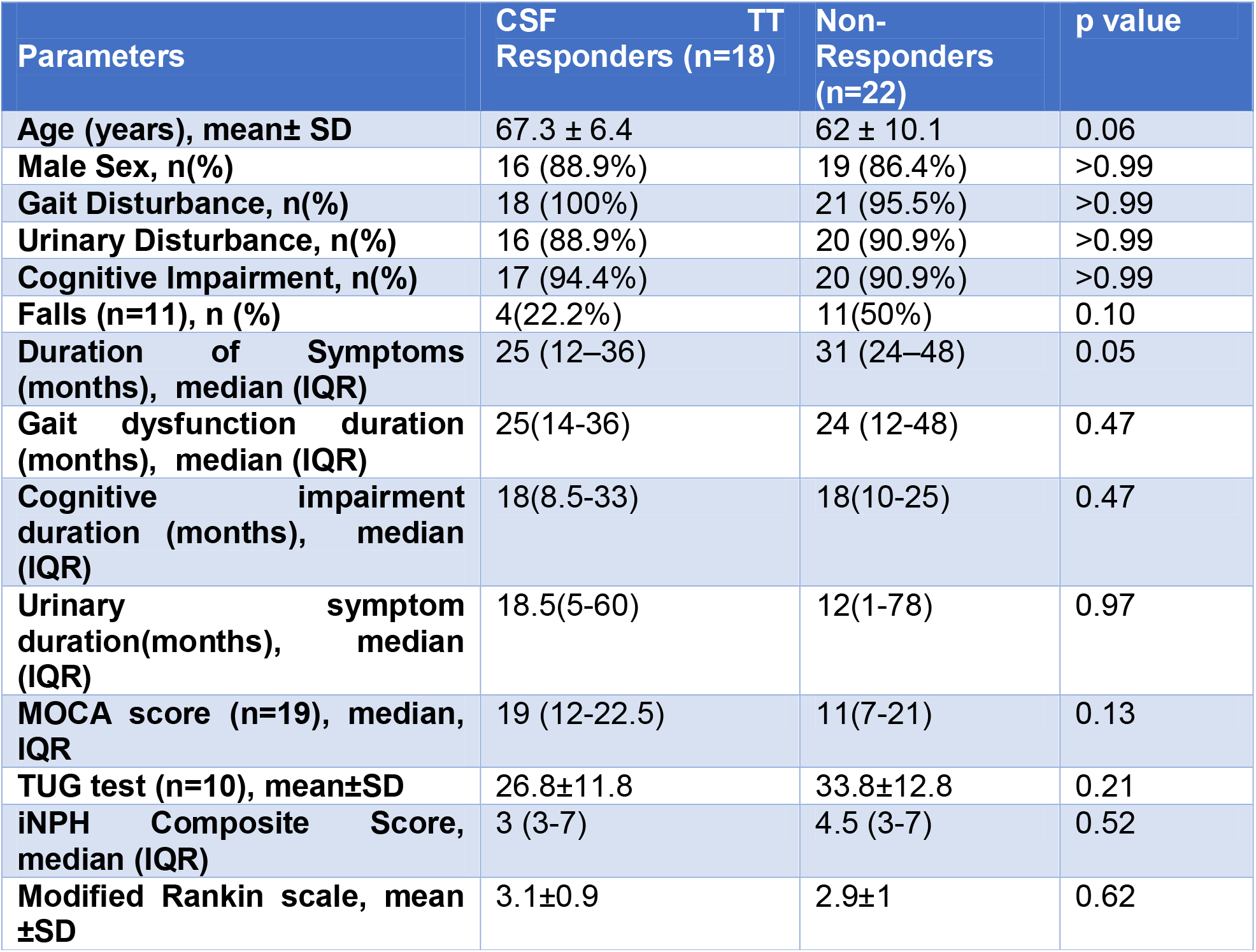
Baseline Characteristics.

### Gait and Functional Outcomes

At baseline, responders and non-responders did not differ significantly in Boon’s gait score, iNPH score, MoCA, TUG, or mRS (Supplementary Table 1). Following the CSF-TT, responders showed significant improvements in select gait subcomponents, particularly in turning disturbance, wide-based stride, and reduced foot clearance. These subscores drove improvements in total walk score and overall Boon’s gait score. MoCA and TUG also improved significantly in responders, while non-responders showed no comparable changes.

### Imaging Findings

MRI parameters—including Evans’ ratio, callosal angle, periventricular white matter changes, DESH, and bowing of the corpus callosum—did not differ significantly between responders and non-responders (Table 2). Imaging features also failed to discriminate shunt responders from non-responders (Supplementary Table 2). Among imaging parameters, none achieved a satisfactory positive predictive value (PPV) for shunt responsiveness, with PPVs ranging from 41% to 69% (Table 3).

**Table 2:**
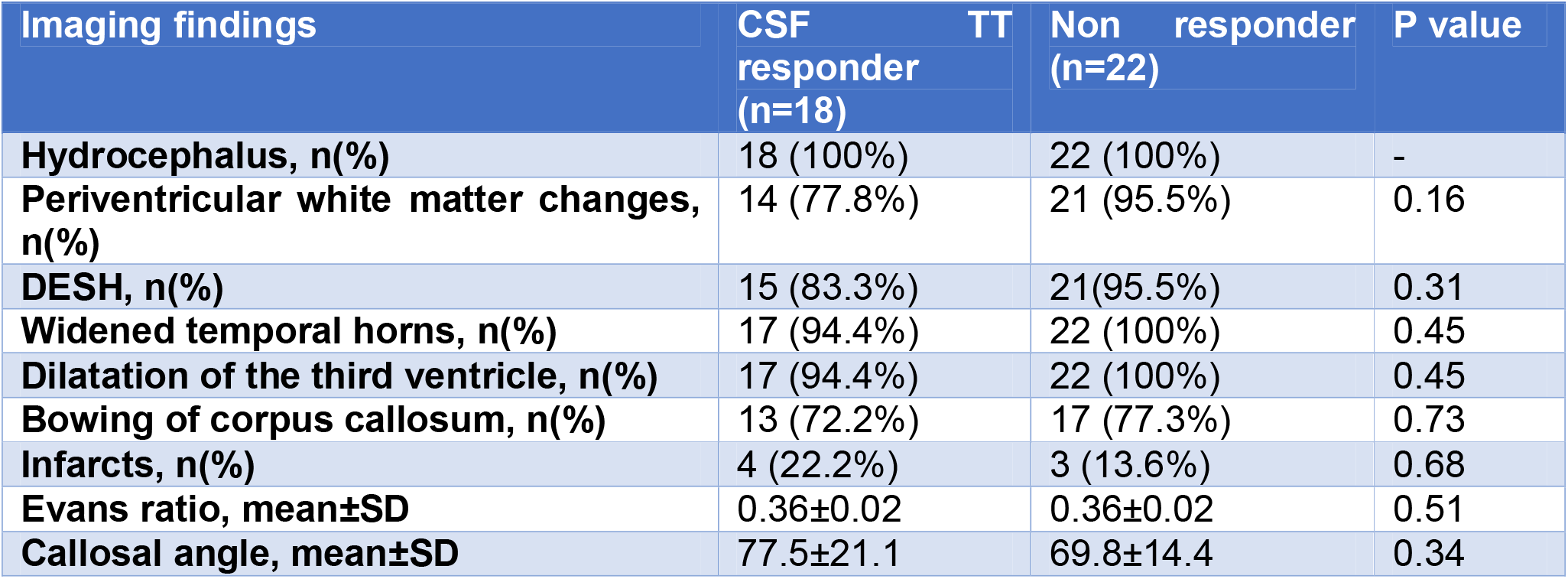
Comparison of imaging findings between CSF TT responders and non-responders.

**Table 3:** Positive predictive values of MRI findings in predicting CSF-TT responsiveness.

|  | PPV for CSF-TT responsiveness (N=24) | PPV for shunt responsiveness (N=40) |
| --- | --- | --- |
| **Periventricular white matter changes** | 40% (23.9% - 57.9%) | 70% (45.7% - 88.1%) |
| **DESH** | 41.7% (25.5% - 59.2%) | 60% (36.1%-80.9%) |
| **Bowing of the corpus callosum** | 43.3% (25.5% - 62.6%) | 68.8% (41.3% - 89%) |
| **Callosal angle $\leq 90$** | 42.4% (25.5% - 60.8%) | 57.9%(33.5%-79.7%) |
| **Any of the above parameters is present** | 45% (30%-61%) | 62.5%(41%- 80%) |

### Composite Diagnostic Accuracy

We tested whether combining CSF-TT responsiveness with elevated CSF opening pressure (greater than 18 cm H_2_O) improves the prediction of shunt response. The composite criterion achieved sensitivity of 62.5% (95% CI: 24.5–91.5) and specificity of 71.4% (95% CI: 29–96), which was modestly higher than CSF-TT alone (sensitivity 53.3%, specificity 66.7%) or opening pressure alone (sensitivity 62.5%, specificity 57.1%) (Table 4). However, confidence intervals were wide, reflecting a limited sample size.

**Table 4:** Diagnostic accuracy of composite diagnostic criteria.

|  | Sensitivity | Specificity | Youden Index |
| --- | --- | --- | --- |
| **To detect shunt responsiveness as defined as at least one point improvement in mRS at 24 weeks post-shunting** |  |  |  |
| **CSF-TT+ opening pressure >18 cm (n=15)** | 62.5% (24.5%-91.5%) | 71.4% (29%-96.3%) | 0.34 |
| **CST-TT (n=24)** | 53.3% (26.6% - 78.7%) | 66.7% (29.9% - 92.5%) | 0.2 |
| **Opening pressure >18 cm H<sub>2</sub>O (n=15)** | 62.5% (24.5% - 91.5%) | 57.1% (18.45-90.15) | 0.2 |

## Discussion

In this Ambispective cohort of patients with probable idiopathic normal pressure hydrocephalus (iNPH), we found that a shorter duration of symptoms was associated with a positive CSF tap test (CSF-TT) response, while other demographic or clinical features did not reliably discriminate responders from non-responders. As anticipated, conventional MRI markers—including DESH, callosal angle, Evans’ ratio, and periventricular changes—did not differ between groups. This is likely because patients were enrolled only if they had imaging features strongly suggestive of iNPH, which limits the discriminative ability of these parameters.

Although ventriculoperitoneal shunt surgery resulted in functional improvement in a substantial proportion of patients^7^, none of the baseline gait features reliably predicted response to CSF-TT or to the surgery. With respect to gait, only three components of Boon’s gait score (turning disturbance, wide-based stride, and reduced foot clearance) showed significant improvement among CSF-TT responders. These subcomponents may therefore be more sensitive indicators of early response than the overall gait scale.

Studies have shown that tight high convexity and medial subarachnoid spaces, as well as enlarged sylvian fissures with ventriculomegaly, defined as DESH, are useful for diagnosing iNPH.^11^ A systematic review reported that only callosal angle and periventricular white matter changes were associated with shunt response, albeit with low diagnostic odds ratios.^12^ Interestingly, while most literature suggests that smaller callosal angles are associated with shunt response, we observed higher angles among shunt responders. This discrepancy may reflect cohort differences or limited sample size. Similar to our study, the study by Alessandra Griffa et al showed that short-term symptom reversal cannot be predicted by either single gait assessment or MRI parameters such as brain morphology, periventricular white matter microstructure, cortical and subcortical perfusion, and white matter lesion load.^13^ DESH, which has been highlighted in Japanese guidelines^3^ as a supportive imaging feature of iNPH, showed a poor positive predictive value of 41.7% in our cohort, while bowing of the corpus callosum demonstrated a PPV of 43%. Our findings align with prior studies that questioned the utility of individual markers.

A multicenter study by Gao et al. showed that combining imaging biomarkers with CSF-TT substantially improved the prediction of shunt responsiveness (AUC 0.72), supporting a multimodal approach.^14^ We also explored composite diagnostic testing by combining CSF-TT responsiveness with elevated CSF opening pressure (>18 cm H_2_O). In this cohort, neither CSF-TT nor opening pressure alone provided high diagnostic accuracy. However, when combined, the sensitivity (62.5%) and specificity (71.4%) improved modestly, suggesting that composite testing may enhance predictive value for shunt responsiveness. These findings should be interpreted with caution, given the wide confidence intervals. Larger studies are needed to validate this approach and to evaluate the role of a composite of modalities such as diffusion tractography, perfusion imaging, CSF biomarkers, and the CSF tap test.

Our study has several strengths. To our knowledge, it is one of the few studies to systematically evaluate the combined role of CSF opening pressure and CSF-TT responsiveness in predicting shunt outcome in iNPH. The ambispective design enabled us to leverage a relatively large cohort from a single tertiary care center with standardized evaluation and follow-up protocols. All patients underwent rigorous clinical, radiological, and CSF assessments based on internationally accepted diagnostic criteria, which strengthens internal validity. The uniform application of MRI parameters and structured gait assessment minimized inter-observer variability. Importantly, the exploration of composite diagnostic markers, rather than single predictors, reflects real-world clinical decision-making and provides a framework for future studies.

Our study has several limitations. First, it was exploratory in nature with a relatively small sample size, and thus not adequately powered to estimate diagnostic accuracy measures with precision. Missing data on CSF opening pressure in the retrospective arm further limited the effective sample size. Second, we relied on single-time-point measurements of both CSF opening pressure and CSF-TT responsiveness. This may have reduced sensitivity and failed to capture fluctuations or delayed improvements that might occur with serial assessments. Third, the ambispective design may have introduced recall bias in the retrospective assessment of postoperative outcomes, which were primarily measured by the modified Rankin Scale; this scale may not fully capture subtler cognitive or urinary changes. Fourth, as a tertiary referral center, our patient population likely represents a more severe spectrum of disease, limiting generalizability. Fifth, we did not include advanced imaging markers such as diffusion tensor imaging, perfusion studies, or CSF biomarker analysis, which could have added diagnostic granularity. Finally, although we explored a composite diagnostic approach, the wide confidence intervals around sensitivity and specificity highlight the need for validation in larger, prospective, multicenter cohorts.

Future research should focus on validating composite diagnostic models that integrate CSF opening pressure, CSF-TT responsiveness, and MRI parameters in larger, prospective, multicenter cohorts. Serial measurements of CSF pressure and repeated assessments of tap-test responsiveness over multiple time points may help capture delayed improvements and refine diagnostic thresholds. Incorporating advanced imaging modalities such as diffusion tensor imaging, tractography, or perfusion studies, along with CSF and blood-based biomarkers, could provide additional pathophysiological insights and improve predictive accuracy. Longitudinal follow-up of patients with “borderline” features, such as those with imaging consistent with iNPH but elevated opening pressures, may help define new clinical subgroups and clarify prognostic differences. Finally, future studies should assess the impact of standardized diagnostic algorithms on surgical decision-making and patient-centered outcomes.

## Conclusion

Shunt surgery remains effective in improving outcomes for carefully selected patients with idiopathic normal pressure hydrocephalus (iNPH). In our study, conventional MRI parameters alone had limited predictive value, while adding CSF opening pressure and CSF tap test responsiveness provided better discriminatory ability to identify shunt responders. These results emphasize the potential of using combined diagnostic methods, although larger prospective studies are necessary to confirm their accuracy and clinical usefulness.

## Data Availability

All data produced in the present work are contained in the manuscript

## Financial Disclosures for the previous 12 months

The authors declare that there are no additional disclosures to report.

## Ethical Compliance Statement

The study was reviewed and approved by the Institute Ethics Committee at the All India Institute of Medical Sciences, New Delhi (No. IECPG-222/20.4.23, RT-14/07.06.23). Informed consent to participate in this study was provided by the participants and their legal guardians. We confirm that we have read the Journal’s position on issues involved in ethical publication and affirm that this work is consistent with those guidelines.

