## Supplementary Table for "Gait parameters, Imaging features, and CSF tap test in idiopathic Normal Pressure Hydrocephalus (iNPH) Is composite testing the way to go?"

Supplementary table 1: Comparison of gait parameters and iNPH scores between CSF TT responders and non responders

| Parameters | PreCSF | | P value | Post CSF | | P value |
| --- | --- | --- | --- | --- | --- | --- |
|  | **CSF TT responder(n=18)** | **CSF TT non responder(n=22)** |  | **CSF TT responder(n=18)** | **CSF TT non responder(n=22)** |  |
| Boon’s gait score | | | | | | |
| Components of walk score | | | | | | |
| *Walking Independently, n(%)* | 15(83.3%) | 16(72.7%) | 0.47 | 17 (94.4%) | 17 (77.3%) | 0.2 |
| *Tandem Walking, n(%)* | 15(100%) | 14(87.5%) | 0.48 | 15 (88.2%) | 14 (82.3%) | >0.99 |
| *Turning Disturbed, n(%)* | 11(73.3%) | 9(56.3%) | 0.46 | 1 (5.88%) | 8 (47.1%) | 0.01 |
| *Trunk Balance, n(%)* | 5(33.3%) | 4(35%) | 0.70 | 5 (29.4%) | 4 (23.5%) | >0.99 |
| *Wide-Based Stride, n(%)* | 7(46.7%) | 8(50%) | >0.99 | 2 (11.8%) | 9 (56.3%) | 0.01 |
| *Small Steps, n(%)* | 11(73.3%) | 12(75%) | >0.99 | 5 (31.3%) | 11 (64.7%) | 0.08 |
| *Reduced Foot Clearance, n(%)* | 12(80%) | 14(87.5%) | 0.65 | 4 (23.5%) | 12 (70.6%) | 0.02 |
| *Start Hesitation, n(%)* | 6(40%) | 6(37.5%) | >0.99 | 1 (5.9%) | 5 (29.4%) | 0.18 |
| *Tendency to Fall, n(%)* | 5(33.3%) | 3(18.8%) | 0.43 | 2 (11.8%) | 4 (23.5%) | 0.66 |
| *Able to Walk assisted, n(%)* | 2(11.1%) | 5(22.7%) | >0.99 | 0 | 4 (18.2%) | 0.33 |
| *Unable to walk at all, n(%)* | 1(5.6%) | 1 (4.5%) |  | 1(5.6%) | 1 (4.5%) |  |
| Total Walk Score, median, IQR | 10 (6-12) | 8 (7-10) |  | 4 (4-4) | 9 (6-16) | <0.01 |
| Steps score, mean±SD | 7.9±2.1 | 7.2±2.7 | 0.39 | 6±2.5 | 6.2±3 | 0.79 |
| Time score mean±SD | 7.9±2.4 | 7.8±2.6 | 0.88 | 5.8±2.5 | 7.1±2.7 | 0.15 |
| Boon’s Gait Scale Total Score mean±SD | 26.1±7.1 | 25.7±9.3 | 0.89 | 16.2±5.4 | 23±10.1 | 0.01 |
| iNPH score | | | | | | |
| *iNPH Gait Score, median(IQR)* | 1(1-1) | 1(1-3) | 0.79 | 1 (0-1) | 1 (1-2) | 0.05 |
| *iNPH Cognition Score, median(IQR)* | 1(1-2) | 1 (1-2) | 0.79 | 1(1-1) | 1(1-3) | 0.24 |
| *iNPH Urinary Score, median(IQR)* | 1(1-3) | 2(1-3) | 0.25 | 1(1-2) | 1(1-3) | 0.11 |
| *iNPH Composite Score, median(IQR)* | 3 (3-7) | 4.5 (3-7) | 0.52 | 2 (2-4) | 4.5 (2-6) | 0.04 |
| MOCA score, median,IQR | 19 (12-22.5) | 11 (7-21) | 0.14 | 22(16.5-23.5) | 12 (10-24) | <0.01 |
| TUG score(n=10) mean±SD | 26.8±11.8 | 33.8±12.8 | 0.21 | 20.4±6.1 | 30.3±15.1 | 0.04 |
| Modified Rankin Score, mean ±SD | 3.1±0.9 | 2.9±1 | 0.62 | 1.89±0.8 | 2.9±1 | <0.01 |
| mRS 1*, n(%)* | 0 | 1 | 0.97 | 6 (33.3%) | 1 (4.6%) | 0.02 |
| mRS 2*, n(%)* | 5(27.8%) | 7 (31.8%) |  | 9 (50%) | 7 (31.8%) |  |
| mRS 3*, n(%)* | 8 (44.4%) | 8 (36.4%) |  | 2 (11.1%) | 8 (36.4%) |  |
| mRS 4*, n(%)* | 4 (22.2%) | 5 (22.7%) |  | 1 (5.6%) | 5 (22.7%) |  |
| mRS 5*, n(%)* | 1 (5.6%) | 1 (4.6%) |  | 0 | 1 (4.6%) |  |

Supplementary Table 2: MRI findings comparison between shunt responders and non-responders

| Imaging findings | shunt responder (n=15) | Non responder (n=9) | P value |
| --- | --- | --- | --- |
| Periventricular white matter changes, n(%) | 14 (93.3%) | 6 (66.7%) | 0.13 |
| DESH, n(%) | 12 (80%) | 8 (88.9%) | >0.99 |
| Widened temporal horns, n(%) | 14 (93.3%) | 9 (100%) | >0.99 |
| Dilatation of third ventricle | 15 (100%) | 9 (100%) | - |
| Bowing of corpus callosum, n(%) | 11 (73.3%) | 5 (55.6%) | 0.41 |
| Infarcts, n(%) | 5 (33.3%) | 2 (22.2%) | 0.67 |
| Evans ratio ,mean±SD | 0.36±0.23 | 0.36±0.20 | 0.51 |
| Callosal angle,mean±SD | 77.5±21.1 | 69.8±14.4 | 0.34 |
